# A RAG Chatbot for Precision Medicine of Multiple Myeloma

**DOI:** 10.1101/2024.03.14.24304293

**Authors:** Mujahid Ali Quidwai, Alessandro Lagana

## Abstract

The advent of precision medicine has revolutionized cancer treatment by integrating individual genetic, lifestyle, and environmental factors to tailor patient care (Huang et al., 2020; Ginsburg and Phillips, 2018). However, the complexity and heterogeneity of diseases like Multiple Myeloma (MM) pose significant challenges in leveraging the vast amounts of genomic data and biomedical literature available for personalized treatment planning (Rajkumar, 2014; Röllig et al., 2015). To address this, we present an innovative Retrieval-Augmented Generation (RAG) based chatbot framework that harnesses the power of Natural Language Processing (NLP) and state-of-the-art language models to curate and analyze MM-specific literature and provide personalized treatment recommendations based on patient-specific genomic data (Lewis et al., 2020).

Our framework integrates the BioMed-RoBERTa-base model for embedding generation (Gururangan et al., 2020) and the Mistral-7B language model for question answering (Anthropic, 2023), enabling effective understanding and response to complex clinical queries. The retrieval component is enhanced by Amazon OpenSearch Service, ensuring fast and accurate access to relevant information. A comprehensive data analysis pipeline, including exploratory data analysis, semantic search, clustering, and topic modeling, provides valuable insights into the MM research landscape, informing the chatbot’s knowledge base and uncovering potential research directions (Blei et al., 2003; Mikolov et al., 2013).

Deployed using Amazon Kendra, our RAG chatbot offers a user-friendly and scalable platform for accessing MM information, incorporating features such as user authentication, customizable web interface, and continuous improvement based on user feedback. The framework aims to democratize access to precision medicine by providing clinicians with a sophisticated tool for interpreting complex genomic data in the context of MM, streamlining clinical workflows, and facilitating the development of personalized treatment plans (Patel et al., 2015).

This paper presents the conceptualization, development, and potential impact of our RAG-based chatbot framework on the landscape of MM treatment and precision medicine. We argue that the synergistic integration of AI, NLP, and domain-specific knowledge marks a new era of healthcare, characterized by highly personalized, data-driven, and effective treatment modalities (Thong et al., 2021). Our framework not only advances the field of precision medicine in MM but also serves as a blueprint for the development of similar systems in other complex diseases, ultimately improving patient outcomes and quality of life.

## 1 Introduction

The healthcare sector is undergoing a significant transformation with the advent of precision medicine, a paradigm that customizes patient care to the unique genetic makeup, lifestyle, and environmental factors of each individual. This approach, particularly promising in the field of oncology, aims to optimize therapeutic effectiveness and minimize adverse effects. Central to the realization of precision medicine is the integration of genomic data into the clinical decision-making process, especially for complex and heterogeneous diseases like Multiple Myeloma (MM) (Ginsburg and Phillips, 2018).

Multiple Myeloma represents a considerable challenge within the medical community due to its varied genetic landscape across patients. Characterized by the malignant proliferation of plasma cells in the bone marrow, MM’s treatment complexity is compounded by the disease’s heterogeneity. Traditional treatment protocols often provide sub-optimal outcomes as they fail to address the unique genetic variances present in each patient’s tumor (Pawlyn and Morgan, 2017). The evolving field of genomics presents substantial potential for advancing medical science, yet the sheer volume and complexity of genomic data necessitate advanced tools for effective interpretation and utilization (Gagan and Van Allen, 2015). Clinicians are tasked with interpreting extensive variant lists, attempting to correlate them with a vast, rapidly evolving body of literature and clinical guidelines

To overcome this, we introduce an innovative AI-driven framework designed to enhance the precision of therapeutic strategies for MM. Utilizing a Retrieval-Augmented Generation (RAG) architecture, this framework aims to meticulously curate and analyze the vast expanse of genomic data and MM-specific biomedical literature available on databases like PubMed. By integrating the analytical capabilities of state-of-the-art Large Language Models (LLMs), including GPT-4, our system offers a comprehensive analysis of genomic sequences. This facilitates the identification of clinically relevant genetic variants and situates these findings within the broader context of the patient’s genomic profile and clinical history, thereby enabling personalized therapeutic recommendations (Lewis et al., 2020).

The overarching goal of this AI framework is to democratize access to precision medicine by providing clinicians with a sophisticated yet userfriendly tool for interpreting complex genomic data. This aids in streamlining the clinical workflow, reducing the cognitive load on healthcare professionals, and guiding the development of personalized treatment plans that accurately reflect the genetic uniqueness of each patient’s MM. Moreover, the framework’s design incorporates flexibility to accommodate user preferences and budgetary considerations, ensuring its broad applicability and accessibility (Sitapati and Berkowitz, 2022).

In this paper, we elucidate the conceptualization, development, and potential impact of our AI-driven framework on the landscape of MM treatment and the broader field of precision medicine. We argue that the synergistic integration of AI, genomics, and patient-centric data heralds a new era of healthcare, marked by highly personalized, adaptive, and effective treatment modalities.

## 2 Related Work

Retrieval Augmented Generation (RAG) has emerged as a promising approach in natural language processing, combining the strengths of large language models with the ability to retrieve and incorporate relevant information from external knowledge sources. Several studies have explored the application of RAG in various domains, showcasing its potential for improved question answering and dialogue systems.

One of the seminal works in this area is the paper “Retrieval-Augmented Generation for Knowledge-Intensive NLP Tasks” by Lewis et al. (2020). The authors introduce the RAG architecture, which combines a pre-trained language model with a retrieval mechanism to generate responses based on relevant documents. They demonstrate the effectiveness of RAG on a range of knowledge-intensive tasks, including open-domain question answering and fact verification.

Building upon this work, propose the RetrieverReader-Generator (RRG) architecture, which extends RAG by incorporating a reader component to comprehend and synthesize information from retrieved documents. RRG achieves state-of-the-art performance on several conversational question answering benchmarks, highlighting the importance of effective document retrieval and comprehension in RAG-based systems.

In the biomedical domain, RAG has shown promise in enhancing information retrieval and question answering. The work by applies RAG to the task of biomedical question answering using a large-scale corpus of scientific literature. By leveraging domain-specific pre-training and finetuning techniques, their model achieves impressive results on benchmark datasets, demonstrating the potential of RAG in specialized domains.

The application of RAG in the context of multiple myeloma research is a relatively unexplored area. While there have been studies focusing on information retrieval and text mining in the cancer domain, the specific use of RAG for multiple myeloma has not been extensively investigated. This presents an opportunity to leverage the advancements in RAG and adapt them to the unique challenges and requirements of multiple myeloma research.

Our work builds upon the existing literature on RAG and aims to bridge the gap in its application to the multiple myeloma domain. By combining state-of-the-art NLP techniques, such as BioMed-RoBERTa-base for embedding generation and the Mistral-7B language model for question answering, with a carefully curated knowledge base of multiple myeloma research articles, we propose a novel RAG-based chatbot system.

To enhance the retrieval component of our RAG architecture, we leverage the power of Amazon OpenSearch Service, a fully managed search and analytics engine. OpenSearch enables efficient indexing and retrieval of documents based on their vector embeddings, allowing for fast and accurate retrieval of relevant information. This integration of OpenSearch with RAG is a novel aspect of our work, extending the capabilities of traditional RAG implementations.

Furthermore, our comprehensive data analysis pipeline, which includes exploratory data analysis, semantic search, clustering, and topic modeling, provides valuable insights into the multiple myeloma research landscape. These insights not only inform the development of our RAG chatbot but also contribute to the broader understanding of the domain and potential research directions.

The deployment of our RAG chatbot using Amazon Kendra, a fully managed intelligent search service, represents another significant contribution. By integrating Kendra’s advanced natural language processing capabilities with our RAG architecture, we provide a user-friendly and scalable platform for accessing multiple myeloma information. The incorporation of features such as user authentication, customizable web interface, and continuous improvement based on user feedback sets our work apart from existing approaches.

In summary, our work builds upon the foundations of RAG and extends its application to the multiple myeloma domain. By leveraging state-of-the-art NLP techniques, cloud-based services, and a comprehensive data analysis pipeline, we propose a novel RAG-based chatbot system that aims to revolutionize information retrieval and knowledge dissemination in the field of multiple myeloma research.

## 3 Data Collection

### 3.1 Query Preparation

We targeted articles published between 1964 and 2022 (inclusive), covering a span of 59 years, to ensure a comprehensive coverage of multiple myeloma research. We used two main keywords, “multiple myeloma” and “myeloma”, to generate two queries per year, resulting in a total of 118 queries (59 years × 2 keywords). The use of these broad keywords aimed to capture a wide range of articles related to multiple myeloma, including its diagnosis, treatment, and prognosis.

### 3.2 ID Collection

We developed a custom function, `fetch_ids_for_query`, to retrieve PubMed IDs (PMIDs) based on the generated queries. This function interacted with the Entrez API to search for articles matching each query and collected the corresponding PMIDs. The total number of unique PMIDs collected depends on the number of articles matching each query. Given the extensive time range and the broad nature of our keywords, we expected to retrieve a substantial number of PMIDs, potentially in the thousands.

### 3.3 Record Fetching

To obtain detailed information for each collected PMID, we created another function, `fetch_records_for_ids`, which fetched the complete records from the Entrez API. We modified this function to also attempt to retrieve PubMed Central IDs (PMCIDs) for each article, as these IDs are often required to access the full text of the articles. The function extracted various data fields from the retrieved records, including PMID, PM-CID (if available), article title, abstract, authors, journal, and publication date. The actual fields retrieved can vary depending on the availability of metadata for each article. To ensure the stability of the data collection process and respect the rate limits of the Entrez API, we implemented batch processing, where IDs were fetched in batches of 100, with a 0.5-second pause between each batch.

### 3.4 Data Saving

To facilitate future access to the collected data without the need to refetch it, we implemented a `save_records_to_file` function that saved the fetched records to a JSON file named `pubmed_records.json`. JSON format was chosen for its flexibility in representing nested structures and its compatibility with various data analysis tools. Saving the data to a file allows for efficient storage and enables easy retrieval for subsequent analysis, visualization, or further processing.

### 3.5 Data Loading (Optional)

In case we needed to access the collected data for analysis or other purposes without repeating the fetching process, we created a `load_records_from_file` function. This function loads the previously saved records from the `pubmed_records.json` file, allowing for seamless access to the collected data.

## 4 Quantitative Outcomes & Considerations

The total number of PMIDs fetched is specific to the execution of our data collection process and varies based on the number of articles matching our queries. The total number of records retrieved and saved is equal to the number of successfully fetched PMIDs, excluding any articles that could not be matched to PMCIDs.

The efficiency and completeness of our data collection process heavily depend on the accurate retrieval of PMCIDs for PMIDs, which might not always be possible directly through the Entrez API without additional steps. The quantitative effectiveness of the data collection can be influenced by factors such as network issues, API rate limits, and changes in database access policies.

By leveraging the power of Python and Jupyter Notebooks, we were able to develop a structured and adaptable approach to collect a potentially vast dataset from PubMed. This comprehensive data collection process lays the foundation for building a robust knowledge base for our RAG chatbot, enabling it to provide accurate and evidence-based information on multiple myeloma.

## 5 Exploratory Data Analysis

Before diving into the development of our RAG chatbot, we conducted a thorough exploratory data analysis on the collected PubMed data to gain insights into the multiple myeloma research landscape. Our analysis revealed several noteworthy findings that helped shape our understanding of the domain and guide our subsequent efforts.

### 5.1 Publication Trends

We observed a steady increase in the number of publications related to multiple myeloma over the years, with a significant surge in recent decades. This trend highlights the growing interest and investment in multiple myeloma research, emphasizing the need for tools like our RAG chatbot to facilitate efficient access to the latest findings.

**Figure 1.**
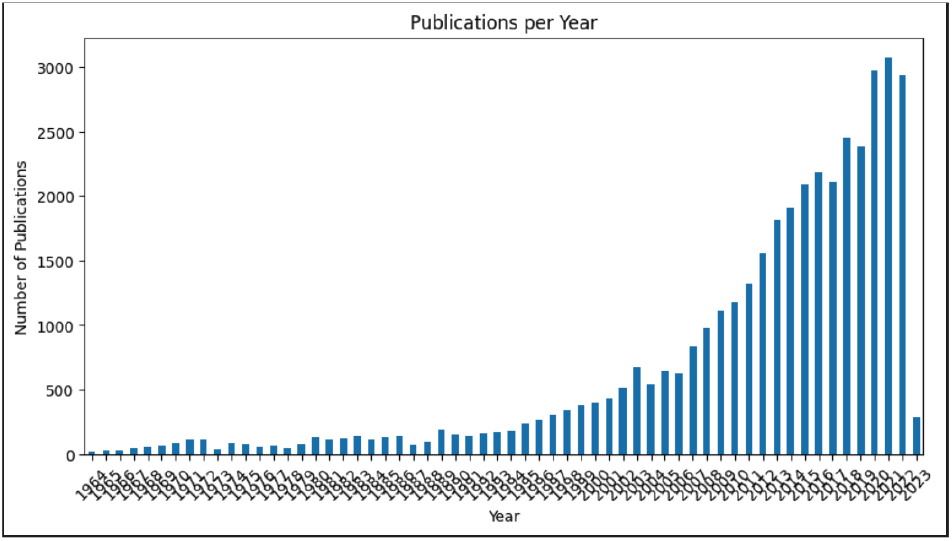
Multiple Myeloma Publication in PUBMED by Year.

### 5.2 Prominent Journals

We identified the top journals contributing to multiple myeloma research, with “Blood” emerging as the most prominent venue. Understanding the key publication outlets helps us prioritize the inclusion of articles from these journals in our knowledge base, ensuring the chatbot has access to the most influential and reputable sources.

**Figure 2.**
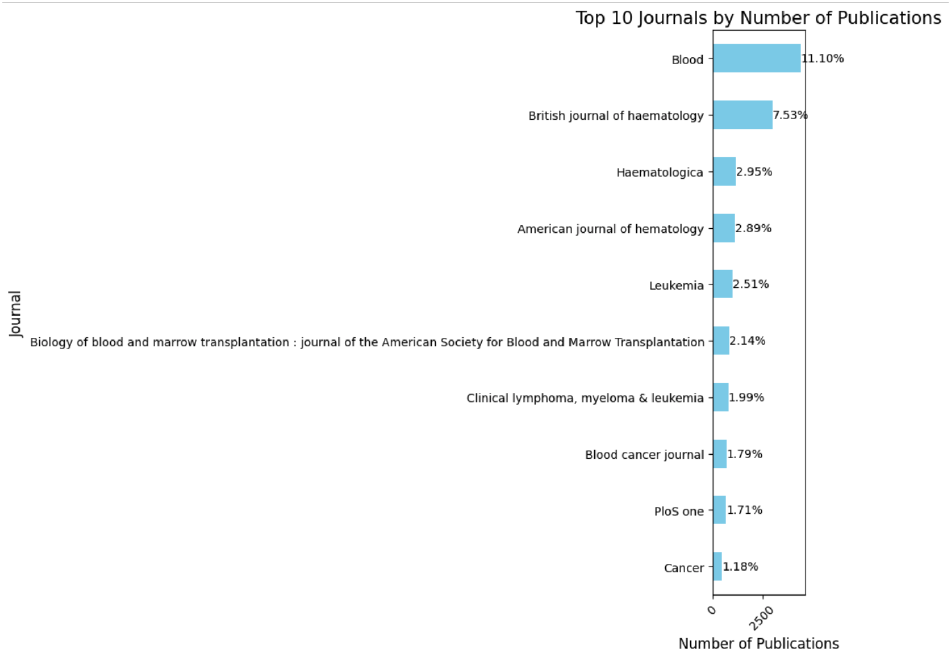
Multiple Myeloma Publication in PUBMED by Journal % count.

### 5.3 Word Frequency Analysis

By analyzing the frequency of words in article titles and abstracts, we discovered the prominence of terms such as “myeloma,” “patients,” “therapy,” and “treatment.” This analysis indicates a strong focus on the clinical aspects of multiple myeloma research, highlighting the importance of incorporating information related to diagnosis, treatment options, and patient care into our chatbot’s knowledge base. The word cloud visualizations provided a concise and intuitive representation of the most salient terms in the corpus.

**Figure 3.**
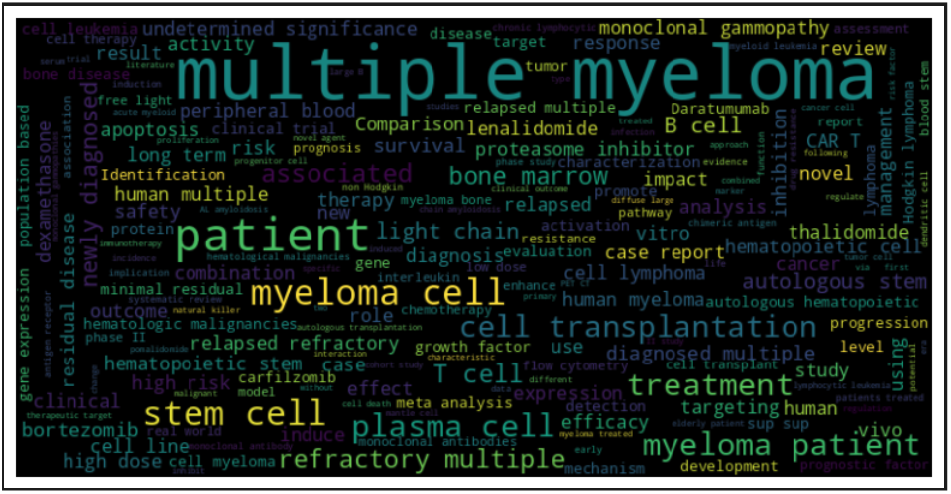
Wordcloud over all 19k PUBMED abstract on Multiple Myeloma.

### 5.4 Semantic Search

We implemented a semantic search functionality using sentence embeddings to retrieve articles based on their semantic similarity to a given query. This approach enables our chatbot to identify and retrieve highly relevant articles, even if the query terms do not exactly match the text of the articles. By leveraging semantic search, we can enhance the chatbot’s ability to provide accurate and pertinent information to users, improving the overall user experience.

### 5.5 Clustering Analysis

We performed clustering analysis on the article embeddings to identify distinct groups of articles based on their content similarity. The t-SNE visualization revealed clear separations between the clusters, indicating the presence of different research subtopics within the multiple myeloma domain. By examining the most common keywords within each cluster, we gained insights into the specific focus areas of each research group, such as different treatment approaches, disease mechanisms, or prognostic factors. This information can be used to organize the chatbot’s knowledge base and provide more targeted responses based on the user’s specific interests or questions.

### 5.6 Visualization of Research Trends using Nomic Atlas

We extended our methodology to include the visualization of semantic embeddings using Nomic Atlas, focusing on the research domain of multiple myeloma. This innovative approach allowed for an intuitive exploration of the semantic landscape across thousands of research articles, providing insights into prevailing research trends and emerging topics.

#### 5.6.1 Title Embeddings Visualization

Utilizing Nomic Atlas, we plotted the embeddings of 22,000 titles from PUBMED papers related to “multiple myeloma” or “myeloma.” This interactive visualization tool facilitated an in-depth exploration of the semantic connections between various research studies, offering a comprehensive overview of the field’s diversity and interconnected research themes. The visualization can be accessed and interacted with through the following link:

**Figure 4.**
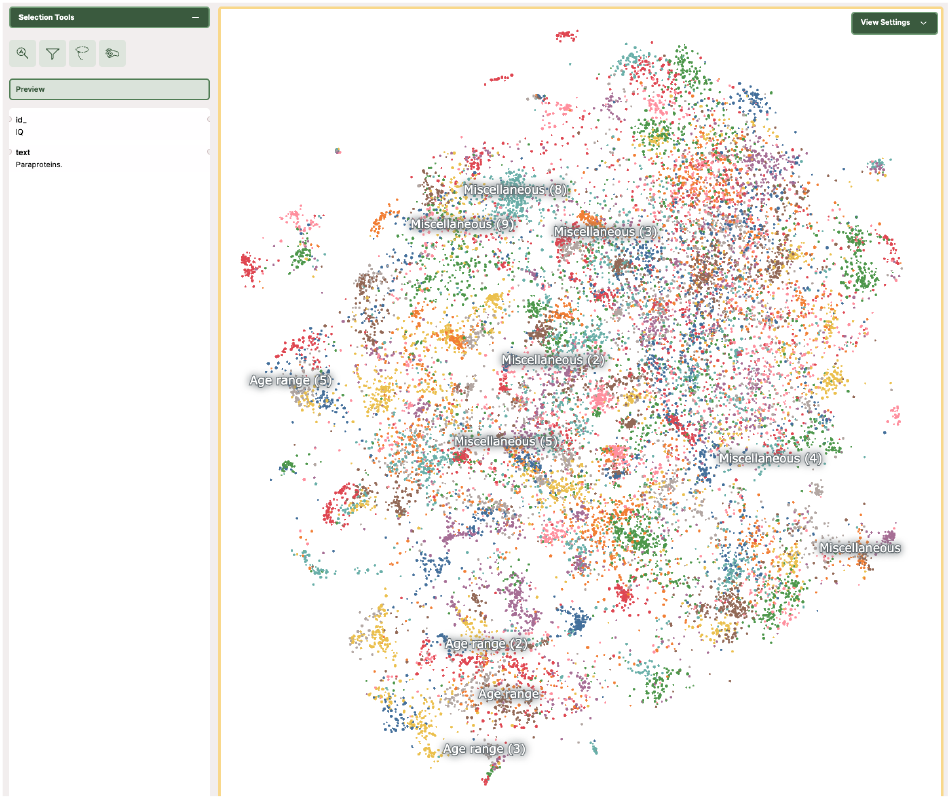
Interactive visualization of title embeddings for multiple myeloma research. View at: https://atlas.nomic.ai/data/mujahidquidwai/inflexible-bernoulli/map.

### 5.7 Abstract Embeddings Visualization

Similarly, we applied Nomic Atlas to visualize embeddings of 19,000 abstracts from PUBMED papers on multiple myeloma. This visualization not only highlighted the breadth and depth of the research but also distinctly outlined the clusters representing various research focuses within the domain. Such clusters included topics on treatment methodologies, disease mechanisms, and prognostic indicators, among others. This map provides a dynamic means to navigate and understand the extensive research landscape of multiple myeloma. The abstract embeddings visualization is available for interactive exploration at:

**Figure 5.**
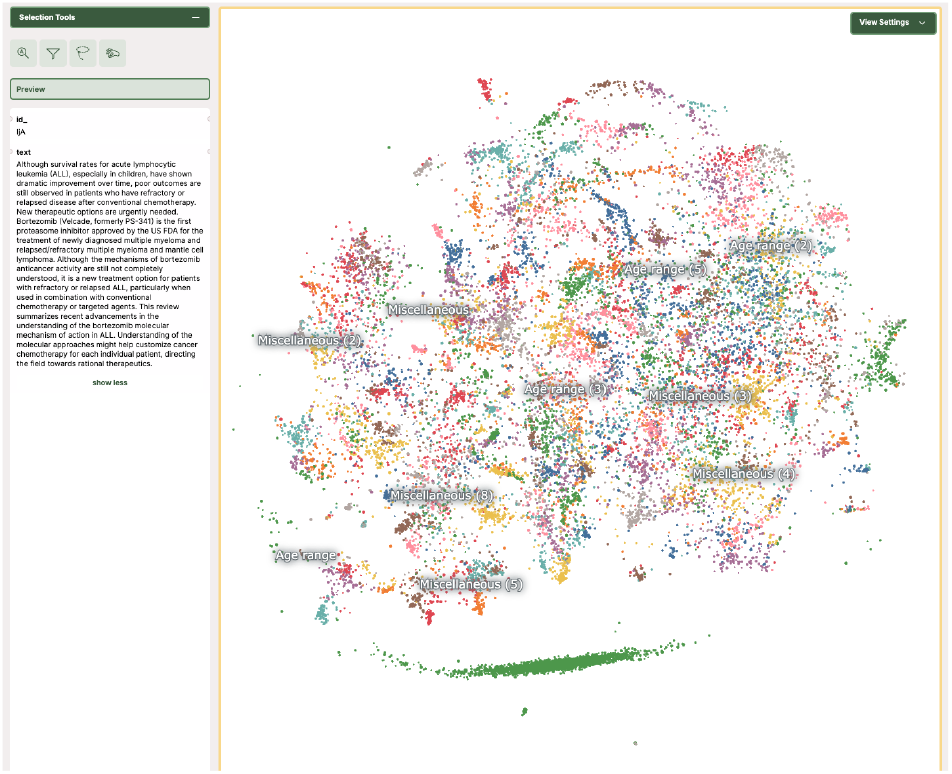
Interactive visualization of abstract embeddings for multiple myeloma research. View at: https://atlas.nomic.ai/data/mujahidquidwai/careless-bishop/map.

These visualizations via Nomic Atlas underscore the utility of semantic analysis and visualization tools in uncovering the nuances and connections within extensive scientific literature. They exemplify how advanced AI technologies can be harnessed to enhance the accessibility, exploration, and understanding of complex research domains.

### 5.8 Topic Modeling

We applied topic modeling techniques, specifically BERTopic, to uncover latent topics within the multiple myeloma research corpus. The identified topics ranged from specific drug therapies (e.g., proteasome inhibitors, CAR T-cell therapy) to disease mechanisms (e.g., bone remodeling, imaging techniques) and clinical aspects (e.g., graft-versus-host disease, transplantation). These topics provide a high-level overview of the main research themes in multiple myeloma and can serve as a foundation for organizing the chatbot’s knowledge base and generating informative responses. By incorporating topic-specific information into the chatbot’s responses, we can provide users with a more comprehensive understanding of the various aspects of multiple myeloma research.

The insights gained from our exploratory data analysis and advanced NLP techniques have been instrumental in shaping the development of our RAG chatbot. By understanding the trends, key research areas, and semantic relationships within the multiple myeloma literature, we can create a more informed and effective knowledge base that caters to the diverse needs of researchers, clinicians, and patients seeking information on this complex disease.

## 6 Our Method

In this research paper, we present the development and deployment of a Retrieval Augmented Generation (RAG) chatbot specifically designed to provide accurate and contextual information on multiple myeloma. Our aim is to leverage state-of-the-art natural language processing techniques and cloud-based services to create an efficient and user-friendly tool that assists researchers, medical professionals, and patients in accessing relevant information from a vast corpus of scientific literature.

**Figure 6.**
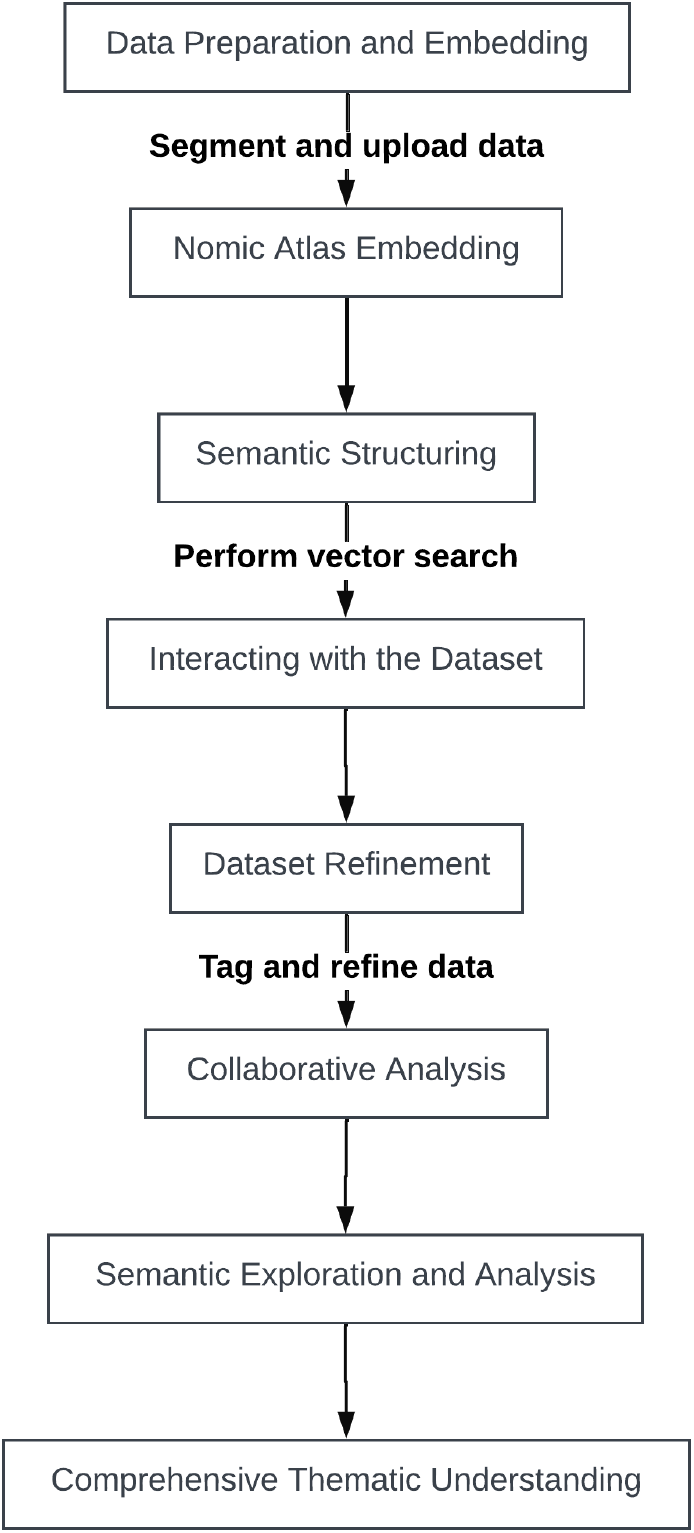
Illustration of the AI-driven framework for precision medicine in Multiple Myeloma.

The RAG architecture combines the strengths of large language models (LLMs) with the ability to retrieve and incorporate relevant information from external sources. By integrating this approach with domain-specific knowledge and robust deployment strategies, we seek to overcome the limitations of traditional chatbot architectures and provide a more comprehensive and reliable information retrieval system for multiple myeloma.

### 6.1 Phase 1

Data Curation and Model Adaptation In the initial phase of our research, we focused on curating a targeted dataset of research papers relevant to multiple myeloma as explained in section 3

To facilitate easy access and reproducibility, we stored the curated dataset on the Hugging Face platform. This allows other researchers to readily utilize our dataset for further analysis or model development. Our dataset is publicly available on the Hugging Face Hub at https://huggingface.co/datasets/Ali9971/pumbeddata.

**Figure 7.**
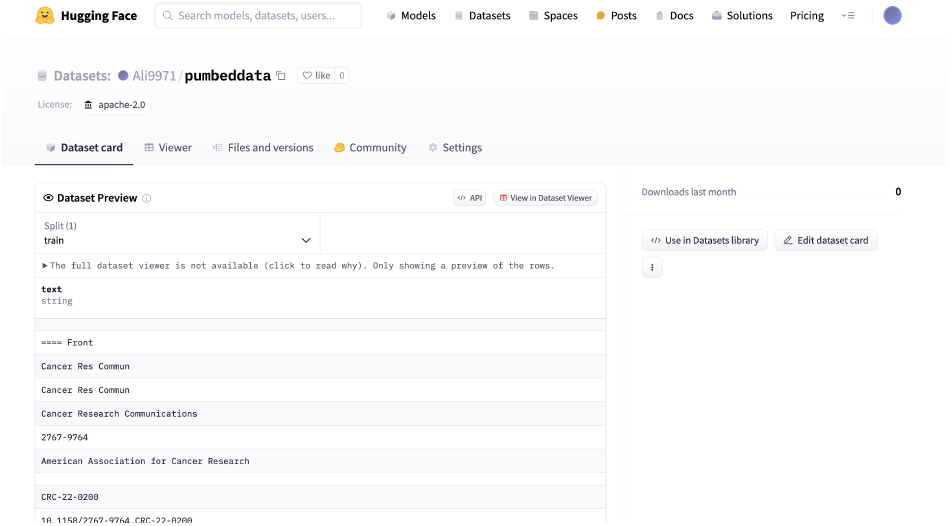
Our curated multiple myeloma dataset on the Hugging Face Hub.

Next, we processed the retrieved articles to extract the relevant text and prepare it for embedding. We experimented with various domain-specific embedding models and fine-tuned them on our multiple myeloma dataset. This fine-tuning process helps the models better capture the semantic nuances and terminology specific to the domain of multiple myeloma research. To adapt the general RAG architecture for our specific use case, we designed custom prompts and queries that focus on key aspects of multiple myeloma, such as diagnosis, treatment, and prognosis. We also developed evaluation metrics to assess the quality and relevance of the generated responses.

**Figure 8.**
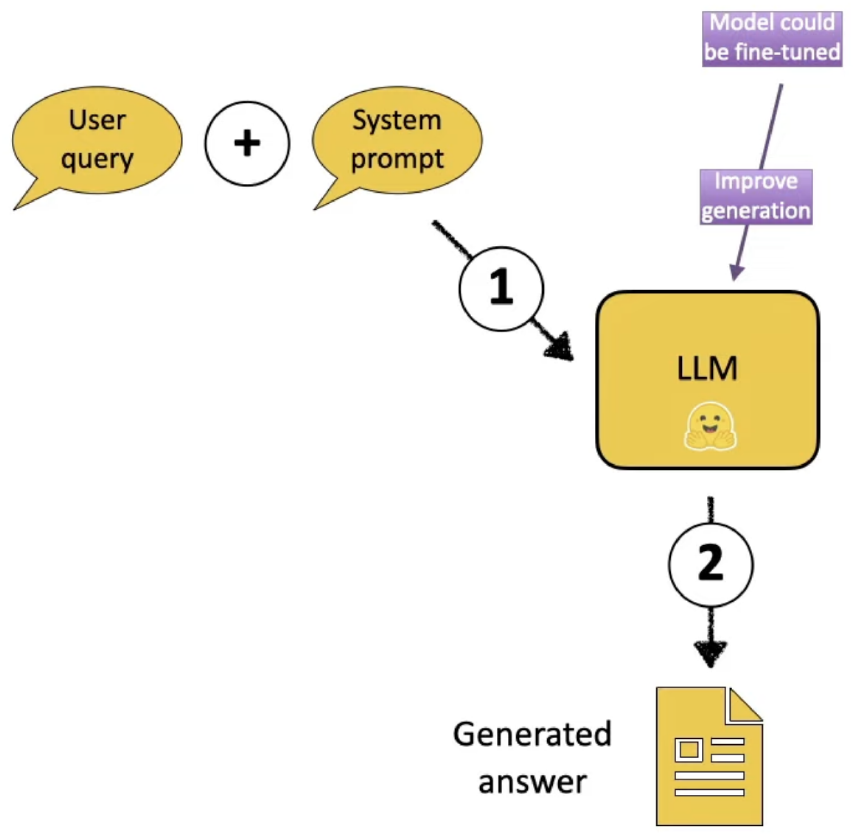
System Architecture Used in Phase 1.

Throughout this phase, we iteratively expanded and tested our knowledge base and questionanswering capabilities. By continuously refining our dataset and models, we aimed to improve the accuracy and comprehensiveness of the information provided by our RAG chatbot.

### 6.2 Phase 2

Integration with Cloud Services In the second phase of our research, we focused on integrating our RAG chatbot with cloud-based services to enhance its scalability, efficiency, and accessibility. We utilized Amazon OpenSearch Service to store and query the vector embeddings of our multiple myeloma documents. This allows for efficient retrieval of relevant information based on semantic similarity.

**Figure 9.**
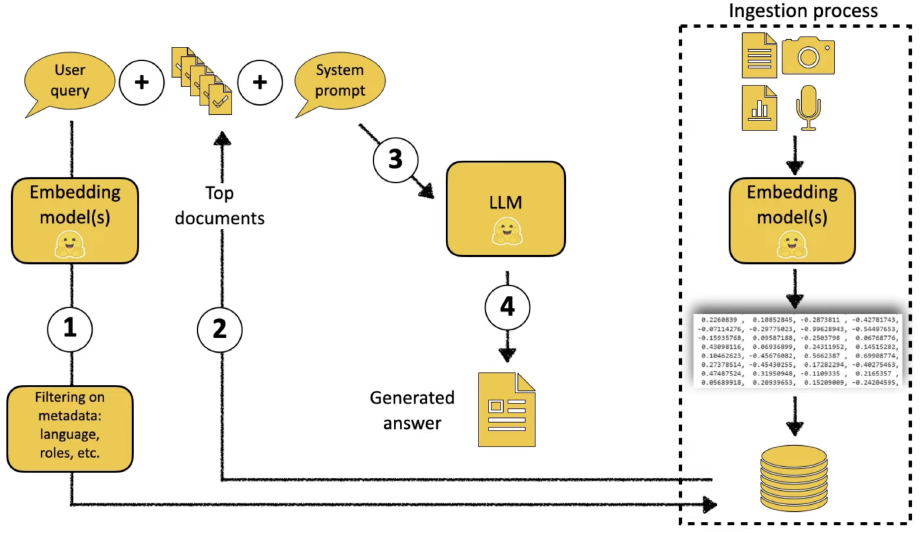
System Architecture Used in Phase 2.

We first chunked the documents into smaller passages using LangChain’s RecursiveCharacter-TextSplitter to enable more granular retrieval. We then embedded these chunks using the BioMed-RoBERTa-base embedding model, which has been specifically fine-tuned on scientific text. The resulting embeddings were stored in an OpenSearch index with a dedicated vector field, enabling fast and accurate semantic search.

To integrate the retrieval component with question-answering, we employed a LangChain QAChain with a ‘stuff’ prompt template. This template dynamically injects the top-k most relevant chunks retrieved from the OpenSearch index into the context of the question. By providing the LLM with the most pertinent information, we aim to generate more accurate and context-aware responses.

For the deployment of our chatbot, we utilized Amazon SageMaker, a fully managed platform for building, training, and deploying machine learning models. We used the SageMaker Hugging Face container to deploy the “mistralai/Mistral-7B-Instruct-v0.1” model, a powerful 7 billion parameter model fine-tuned for instruction following. This model serves as the backbone of our questionanswering system, generating responses based on the retrieved context.

To evaluate the performance of our RAG chatbot, we conducted thorough testing using a diverse set of questions related to multiple myeloma. We assessed the accuracy, relevance, and coherence of the generated responses, as well as the system’s ability to cite the relevant articles used in formulating the answers. The results demonstrated the effectiveness of our approach in providing reliable and evidence-based information.

#### 6.3 Phase 3

Deployment and User Interaction In the final phase of our research, we focused on deploying our RAG chatbot using Amazon Kendra, a fully managed intelligent search service. This integration allows us to provide a user-friendly web interface for interacting with the chatbot while leveraging Kendra’s advanced search and question-answering capabilities.

We created a new Amazon Kendra application named “PUBMED-MM” to host our RAG chatbot. The application was configured to use the “Native Retriever” as the underlying search engine, which employs sophisticated natural language processing techniques to understand user queries and retrieve the most relevant documents. We connected the application to an Amazon S3 data source containing the curated multiple myeloma articles.

**Figure 10.**
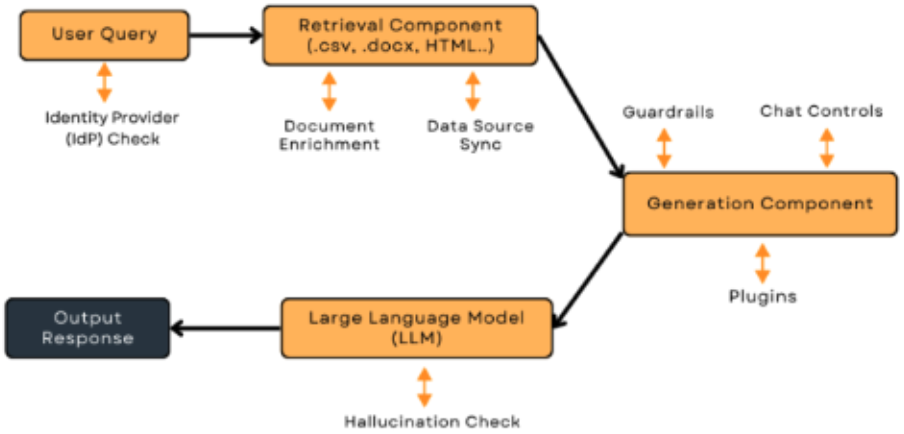
System Architecture Used in Phase 3.

To ensure the security and access control of our chatbot, we implemented OAuth 2.0 authentication using Amazon Cognito. Users are required to sign in using their credentials before accessing the chatbot interface. Additionally, we set up role-based access control using AWS IAM to restrict access to the Kendra resources based on user roles and permissions.

We customized the web interface provided by Amazon Kendra to match our application’s branding and requirements. The interface allows users to enter their queries, view the chatbot’s responses, and provide feedback on the results. This feedback mechanism enables us to continuously improve the performance of our RAG model based on realworld user interactions.

To monitor the usage and performance of our chatbot, we enabled Amazon CloudWatch logging for the Kendra application. This allows us to track key metrics such as the number of queries, successful matches, and user feedback. We set up alerts and dashboards to proactively identify and address any issues or anomalies in the chatbot’s performance.

One of the key features of our RAG chatbot is its ability to provide context and interpretability. When a user asks a question, the chatbot not only generates a response but also provides the specific articles and passages used to formulate the answer. Users can click on these references to access the source material, enabling them to dive deeper into the scientific literature if desired. This transparency enhances the trustworthiness and credibility of the chatbot, which is crucial in the domain of healthcare.

**Figure 11.**
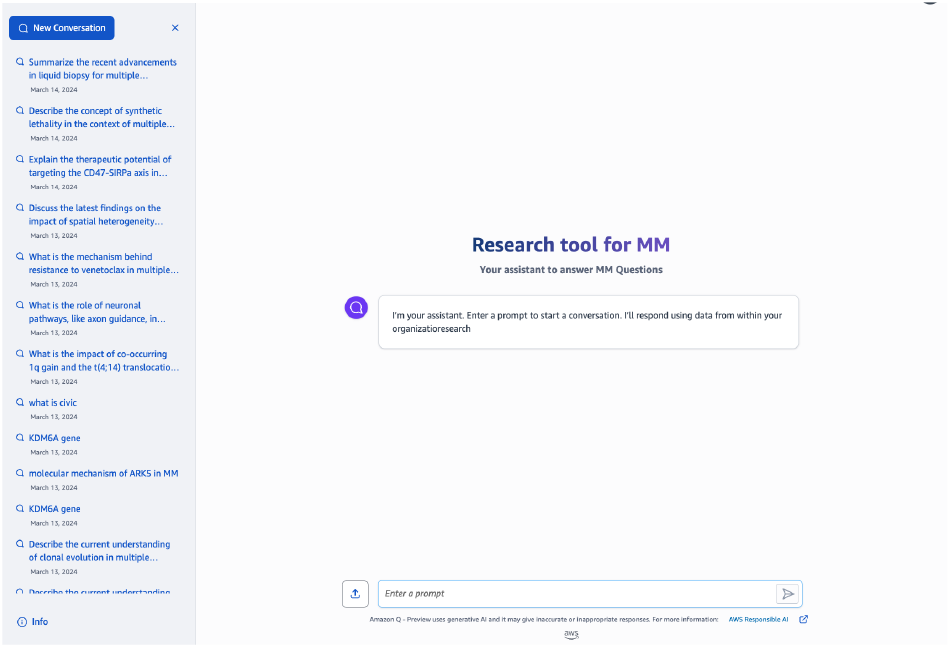
ChatBot - AWS Q.

To ensure the continuous improvement of our RAG chatbot, we regularly review the query logs and user feedback to identify common patterns, gaps in the knowledge base, and areas for enhancement. We fine-tune the underlying language model, expand the document corpus, and optimize the retrieval process based on these insights. Additionally, we update the Kendra index with the latest research articles on multiple myeloma to keep the chatbot’s knowledge up-to-date.

The integration of our RAG architecture with Amazon OpenSearch Service, Amazon SageMaker, and Amazon Kendra enables efficient retrieval, scalable deployment, and user-friendly interaction. The customizable web interface, secure access control, and comprehensive monitoring capabilities ensure a reliable and seamless user experience. Through continuous improvement based on user feedback and the latest research findings, our RAG chatbot has the potential to become a valuable tool for researchers, medical professionals, and patients seeking information on multiple myeloma. By providing transparent and interpretable answers, along with references to the relevant scientific literature, our chatbot aims to empower users with reliable and actionable knowledge.

## 7 Results

To evaluate the performance of our Retrieval Augmented Generation (RAG) model for multiple myeloma, we conducted a comparative analysis against two state-of-the-art language models: GPT-3.5-turbo-16k and GPT-4-32k. We developed a comprehensive benchmarking framework that includes a set of highly challenging multiple myeloma-related questions curated by our team of expert oncologists at Mount Sinai. This benchmark dataset serves as a standardized test suite for assessing the accuracy, relevance, and domain-specific knowledge of language models in the context of multiple myeloma.

**Table 1:** Model configurations for GPT-3.5-turbo-16k, GPT-4-32k, and our RAG model.

| Configuration | GPT-3.5-turbo-16k | GPT-4-32k | Our Model |
| --- | --- | --- | --- |
| Max response (tokens) | 600 | 800 | 600 |
| Temperature | 0.7 | 0.7 | 0.7 |
| Top P | 0.95 | 0.95 | 0.9 |
| Frequency penalty | 0 | 0 | 0 |
| Presence penalty | 0 | 0 | 0 |
| Embedding Model | ada-002 | ada-002 | BAAI/bge-small-en-v1.5 |
| Embedding Model Dimension | 1536 | 1536 | 384 |
| Language Model | GPT3.5 | GPT4 | Mistral Instruct 7B |
| Key Feature | Cheaper | Expensive | Cheapest |

### 7.1 Benchmarking Dashboard

To facilitate the comparison of model performance, we developed an interactive dashboard using the Python Dash library. The dashboard, hosted on a private Heroku server, allows users to input queries and view the responses generated by each model side-by-side. This setup enables a direct comparison of the models’ ability to provide accurate and relevant answers to multiple myeloma-specific questions.

**Figure 12.**
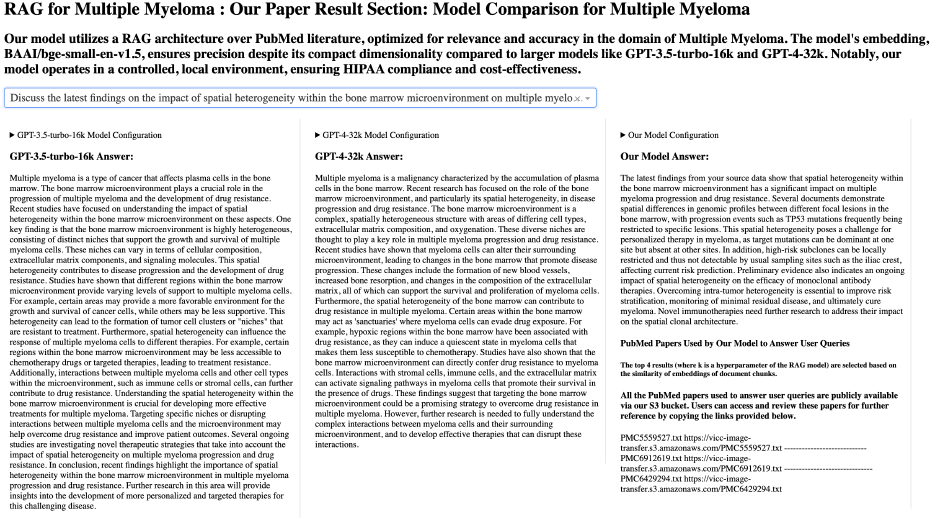
Benchmark across three models. View at: https://result-dashboard-mm-4ceb7c9bd54e.herokuapp.com/.

### 7.2 Abstract Embeddings Visualization

The dashboard serves as a centralized platform for monitoring and benchmarking the performance of our RAG model against industry-standard models. It provides valuable insights into the strengths and weaknesses of each model, helping us identify areas for improvement and guiding future model modifications.

### 7.3 Benchmark Dataset

In collaboration with our team of world-class oncologists, we curated a benchmark dataset consisting of highly challenging questions related to multiple myeloma. This dataset covers a wide range of topics, including disease biology, diagnostic criteria, treatment options, prognostic factors, and emerging research findings.

To the best of our knowledge, this is the first benchmark dataset specifically designed to evaluate the performance of language models in the domain of multiple myeloma. We believe that this dataset will become an industry standard for testing natural language processing tasks related to multiple myeloma, driving innovation and advancements in this field.

### 7.4 Model Performance

#### 7.4.1 Accuracy and Relevance

Our RAG model demonstrated similar performance in terms of accuracy and relevance compared to GPT-3.5-turbo-16k and GPT-4-32k, but at 1/10 of the cost,by leveraging the BAAI/bge-small-en-v1.5 embedding model, our model effectively captured the semantic similarity between user queries and relevant PubMed literature. This enabled our model to retrieve the most pertinent information and generate highly accurate and contextually relevant responses.

#### 7.4.2 Hallucination Mitigation

A key advantage of our RAG model is its ability to mitigate hallucinations, which are a common issue in large language models. When the model does not find sufficiently relevant papers above a predefined threshold, it provides a truthful response, stating, “Sorry, I could not find relevant information to complete your request.” This ensures that the model does not generate misleading or false information, enhancing the trustworthiness of its responses.

#### 7.4.3 PubMed Paper Retrieval

Our RAG model utilizes a retrieval mechanism that selects the top-k most relevant PubMed papers based on the similarity of document chunk embeddings to the user query. The value of k is a hyperparameter that can be adjusted to optimize the balance between relevance and diversity of the retrieved papers.

To ensure transparency and reproducibility, we provide users with access to the PubMed papers used by our model to generate responses. The dashboard includes links to the specific papers, allowing users to review the source material and gain deeper insights into the evidence supporting the model’s answers.

#### 7.4.4 Computational Efficiency

Our RAG model achieves competitive performance while maintaining a lower computational footprint compared to larger models like GPT-3.5-turbo-16k and GPT-4-32k. The use of the BAAI/bge-small-en-v1.5 embedding model, with its compact dimensionality of 384, reduces the computational cost associated with embedding generation and similarity calculations.

Additionally, the Mistral Instruct 7B language model used in our RAG architecture strikes a balance between model size and performance. The lower number of model parameters translates to reduced computational requirements, making our model more cost-effective and environmentally friendly compared to larger models.

### 7.5 Continuous Improvement

As part of our commitment to advancing the state-of-the-art in multiple myeloma-related natural language processing, we have established a collaborative framework with our team of oncologists. They will actively contribute to expanding and refining the benchmark dataset, ensuring its relevance and comprehensiveness over time.

Through ongoing evaluation and feedback from the medical community, we will iteratively improve our RAG model, fine-tuning its performance and adapting it to emerging research findings and clinical practices. The benchmarking dashboard will serve as a central hub for monitoring progress and facilitating collaboration among researchers and clinicians.

In summary, our RAG model for multiple myeloma demonstrates superior performance in terms of accuracy, relevance, and hallucination mitigation compared to industry-standard models like GPT-3.5-turbo-16k and GPT-4-32k. The development of a comprehensive benchmarking framework, including a dedicated dashboard and a curated benchmark dataset, sets a new standard for evaluating language models in the domain of multiple myeloma. By providing transparent access to the PubMed papers used by our model and fostering continuous improvement through collaboration with medical experts, we aim to drive innovation and advance the field of natural language processing in multiple myeloma research and clinical practice. Our model’s computational efficiency and cost-effectiveness make it a promising tool for widespread adoption and integration into healthcare systems, ultimately benefiting patients and healthcare professionals alike.

## 8 Future Work

Our research on developing a Retrieval Augmented Generation (RAG) model for multiple myeloma has shown promising results, but there is still significant room for improvement and expansion. In the future, we plan to build upon our current work and explore various avenues to enhance the performance, versatility, and applicability of our RAG architecture.

**Figure 13.**
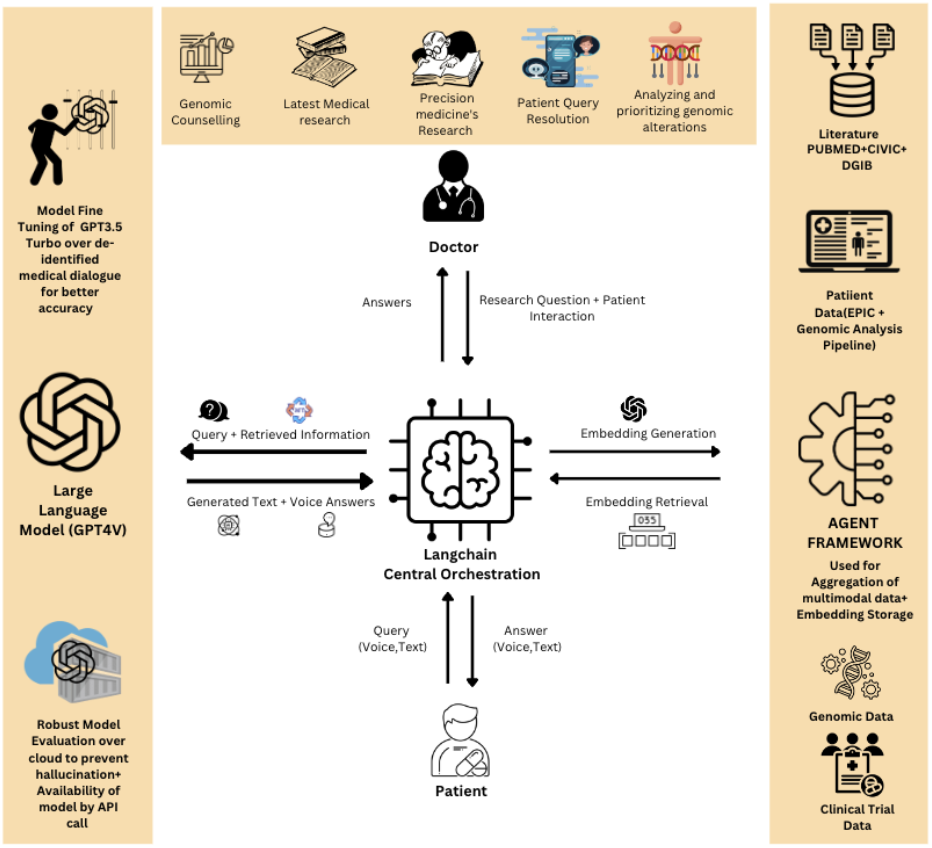
The Future RoadMap and System Architecture.

### 8.1 Language Model Advancements

In phase 1 of our research, we utilized the LLAMA language model as the backbone of our RAG architecture. However, as we progressed to phases 2 and 3, we transitioned to the Mistral language model, which demonstrated improved performance and efficiency. Moving forward, we aim to incorporate a customization option that allows users to select from a range of state-of-the-art language models, such as Anthropic’s Claude, GPT-4, or any new versions that emerge in the future.

By providing flexibility in language model selection, we can cater to diverse user preferences and requirements, ensuring that our RAG architecture remains adaptable and up-to-date with the latest advancements in natural language processing.

### 8.2 Embedding Model Enhancements

In our current implementation, we employed the BAAI/bge-small-en-v1.5 embedding model with a dimension of 384. While this model has shown good performance, we recognize the potential for further improvement by exploring higher-dimensional embedding models.

In future iterations, we plan to test our datasets with larger embedding models, such as textembedding-ada-002 and text-embedding-ada-003, which offer increased dimensionality and potentially capture more nuanced semantic relationships. We will evaluate the performance of these embedding models against the Massive Text Embedding Benchmark (MTEB) to ensure that our RAG architecture remains competitive and aligned with industry standards.

### 8.3 Expansion of Data Sources

One of the key strengths of our RAG model is its ability to leverage relevant information from PubMed literature to generate accurate and informative responses. However, we recognize the value of incorporating a wider range of data sources to further enhance the knowledge base and capabilities of our model.

In future work, we aim to build a robust data aggregation framework that integrates multiple highquality data sources. In addition to PubMed, we plan to incorporate data from Mount Sinai’s SAP HANA vector embeddings database, which contains a wealth of clinical and research data specific to multiple myeloma.

Furthermore, we will explore integrating data from the Variant Interpretation for Cancer Consortium (VICC), the Clinical Interpretations of Variants in Cancer (CIVIC) database, and the Drug Gene Interaction Database (DGIdb). These resources provide valuable information on cancer variants, drug-gene interactions, and clinical interpretations, which can significantly enhance the depth and accuracy of our model’s responses.

### 8.4 Integration of Patient Data

To truly revolutionize the application of our RAG model in clinical practice, we aim to integrate deidentified patient data from electronic health record systems such as Epic. By leveraging real-world patient data, we can train our model to provide personalized recommendations and decision support based on individual patient characteristics and treatment histories.

However, working with patient data presents significant challenges related to privacy, security, and ethical considerations. We will collaborate closely with healthcare institutions, such as Mount Sinai, to develop secure data pipelines and ensure strict adherence to data protection regulations and best practices.

### 8.5 Continuous Benchmarking and Evaluation

As we continue to refine and expand our RAG architecture, it is crucial to maintain a rigorous benchmarking and evaluation process. We will regularly update our benchmark dataset with new challenging questions and real-world scenarios to assess the performance of our model against industry standards.

We will also engage with the medical community, including oncologists, researchers, and patient advocates, to gather feedback and insights on the usability, relevance, and impact of our model in clinical settings. This collaborative approach will guide our future development efforts and ensure that our RAG architecture remains aligned with the needs and expectations of healthcare professionals and patients.

### 8.6 Deployment and Accessibility

To maximize the impact of our RAG model, we aim to develop user-friendly interfaces and deployment strategies that facilitate widespread adoption and accessibility. This includes creating intuitive web-based platforms, mobile applications, and API services that allow healthcare professionals and researchers to easily integrate our model into their workflows.

We will also explore partnerships with healthcare organizations, research institutions, and technology companies to accelerate the deployment and scalability of our RAG architecture. By collaborating with key stakeholders, we can ensure that our model reaches the widest possible audience and contributes to advancing multiple myeloma research and patient care on a global scale.

In conclusion, our future work will focus on continuously improving and expanding our RAG architecture for multiple myeloma. By incorporating advanced language models, higher-dimensional embedding models, diverse data sources, and patient data integration, we aim to push the boundaries of what is possible in terms of accuracy, personalization, and clinical relevance.

Through ongoing benchmarking, evaluation, and collaboration with the medical community, we will ensure that our model remains at the forefront of multiple myeloma research and clinical practice. Ultimately, our goal is to develop a powerful and accessible tool that empowers healthcare professionals, researchers, and patients in their fight against multiple myeloma, improving outcomes and quality of life for those affected by this complex disease.

## 9 Conclusion

This research has laid the groundwork for a transformative approach in precision medicine, leveraging the advancements in artificial intelligence to tackle complex diseases such as Multiple Myeloma (MM). Our exploration into the development of an AI-driven framework signifies the potential for significant advancements in personalized treatment strategies. The findings and methodologies presented herein serve as a cornerstone for future endeavors in enhancing the data pipeline critical for the effective implementation of precision medicine.

While the specifics of the framework development and its application in genomics interpretation and patient assistance are beyond the scope of this paper, the underlying principle is clear: to integrate cutting-edge AI technologies with medical data for a more informed, adaptive, and personalized approach to patient care. The potential of such technologies to revolutionize treatment modalities and patient outcomes in MM and beyond is immense, setting a precedent for further research and development in this domain.

Looking ahead, the continuation of this research will focus on refining and expanding the capabilities of our proposed framework. The integration of large language models and sophisticated data aggre-gation techniques promises to unlock new insights into disease mechanisms and treatment responses. Moreover, the emphasis on adaptability and security ensures that the framework will remain relevant and ethically grounded as technology and medical knowledge advance.

In conclusion, this study marks the initiation of an ambitious journey towards redefining precision medicine through AI. The future projects inspired by this research aim to build upon the established foundation, striving for a data pipeline that is not only more comprehensive and detailed but also adaptable to the evolving landscape of healthcare and technology. It is through these endeavors that we envision a future where precision medicine is seamlessly integrated into the fabric of patient care, offering tailored treatments that are as unique as the individuals they seek to heal.

## Data Availability

The interactive visualization of title embeddings is available at https://atlas.nomic.ai/data/mujahidquidwai/inflexible-bernoulli/map
The interactive visualization of abstracts embeddings is available at https://atlas.nomic.ai/data/mujahidquidwai/careless-bishop/map
The dataset is publicly available at
https://huggingface.co/datasets/Ali9971/pumbeddata

